# Giving Voice to Patients Through Interviews and Qualitative Analysis- A Pilot Study

**DOI:** 10.1101/2020.03.16.20030684

**Authors:** Matthew H Loxton, Ebele Okoli

## Abstract

Modern healthcare is drowning in data, and burdened by quality, safety, financial, and operational metrics, but few relate directly to how patients experiences their care. The literature lacks sufficient specificity on how care processes are seen through the eyes of the patient.

To fill a gap in awareness of the patient experience of the radiology processes, we used a multimethod qualitative approach to elicit the patient view of their radiology experiences and insights from the patient voice. We developed a typology of patient experiences of the radiology processes that centered on communication gaps, and reflected opacity, fragility, and unpredictability of administrative and care processes in radiology that failed to interconnect efficiently or effectively, and did not work well as an end-to-end patient journey. Care processes were described by participants as fragile, solitary, and opaque, and required constant vigilance, supervision, and assistance by patients. Participants described a need for improved communication between radiology staff and patients that focuses on the patient journey and helps to identify and mitigate causes of process opacity and fragility.

## Introduction

One of the key recommendations of the National Patient Safety Foundation (NPSF) is to “create a common set of safety metrics that reflect meaningful outcomes.” [1] This paper attempts to look at patient outcomes from the broad perspective of how patients might experience the end-to-end journey through radiology, and generate a typology of potential measures of quality.

From a quality improvement perspective, patients are the “customers” for radiology care, and their perspective is valuable in improving radiology processes. In this qualitative study, we used a contextual approach to examine radiology from the patient perspective, report on the patient view of their radiology experiences, and attempt to develop a typology.

The interviews described radiology care processes as often opaque, fragile, or extremely siloed, and frequently failed in ways that undermined patient confidence, and caused anxiety or alarm.

**Figure 1.**
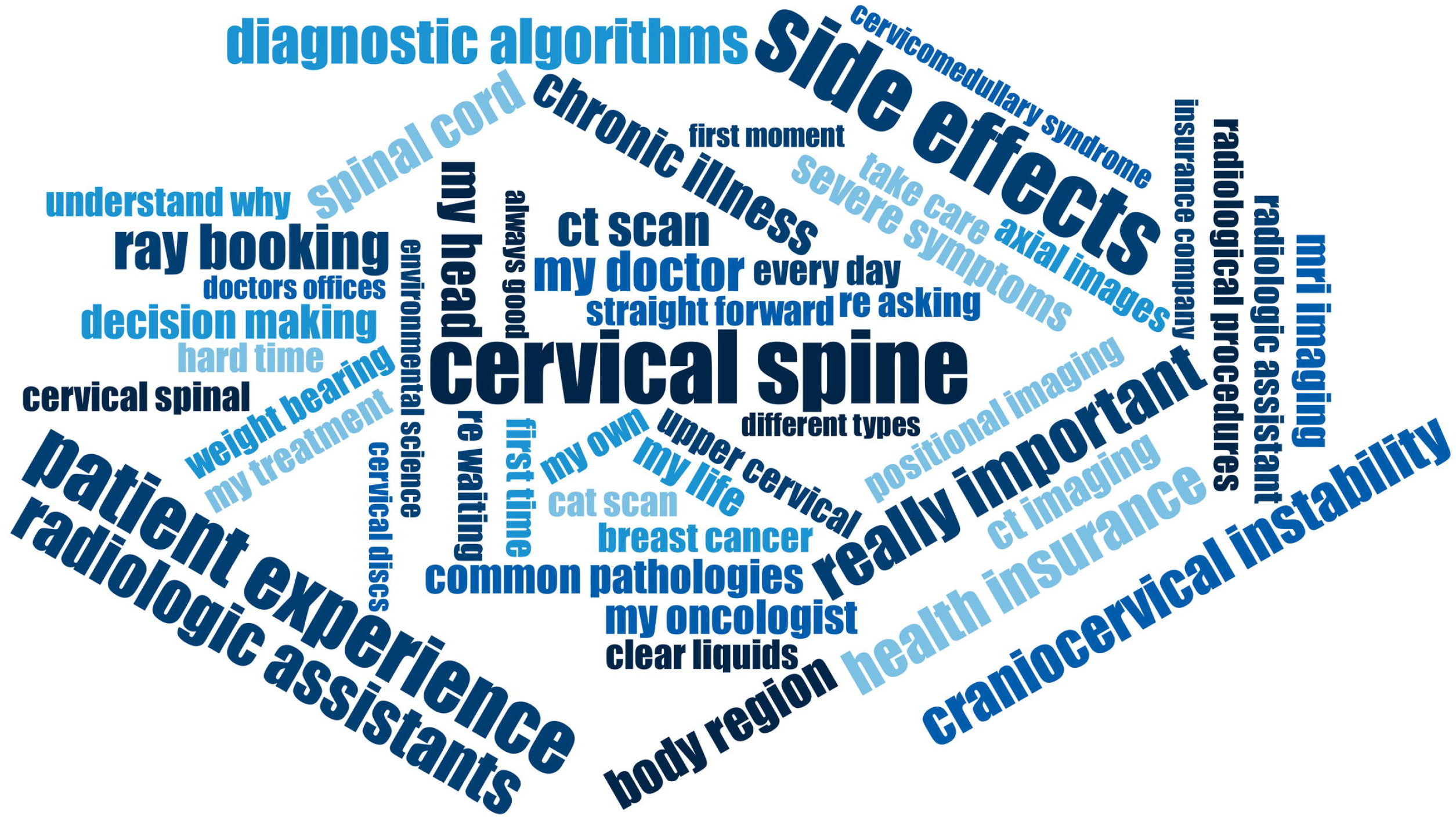
Patient Voice

## Method

This study was evaluated by the corporate ethics team as IRB exempt, and interviewing, coding, and analysis was carried out by two researchers, both with experience in monitorting & evaluation and process improvement, and experience with patient-facing health policy implementation, technology deployment, and workflow improvement assessments. The study uses multi-method [3] and draws from traditions of postpositivism (an external reality), constructivism (participants form own views of healthcare), phenomenalism (how participants experienced healthcare), and grounded theory (coding built mainly from what participants revealed). We did so to establish an asymetrical but “conversational partnership” in which the researchers and participant jointly shaped the path of discovery. [4]

We recruited participants through social media, including Twitter. We invited patients and carers who had previous experience of radiology in a message on Twitter in which we described the aim of the study, and referred interested parties to a Physician’s Weekly Blog entitled “Micro-Study: Discovering the Patient Voice.” Prospective participants were invited to contact the lead researcher by direct message on Twitter, email, or through LinkedIn. Ten individuals made contact, and expressed the wish to be interviewed, and were emailed details outlining the purpose of the study, topic areas, the level of privacy they should expect, and an offer of a $25 gift card for participation. Of the ten respondents, one was lost to contact, and nine completed the study. Participant identity was not verified, and we did not verify experiences or events that participants described.

The use of online patients is an important limitation of this study, as it is difficult to verify their radiology experience, and also limits respondents to those patients and carers who have internet access and are active on social media. Since this is a qualitative study, it seeks to answer the question “what can happen” not “what is typical,” and therefore does not attempt to have a large or representational respondent panel. Participants were interviewed over the phone, and sessions were recorded with explicit participant permission. The interviews were transcribed using an online transcription service with a machine learning system, and was coded in a qualitative data analysis (QDA) tool. The transcription results were manually compared to the audio track.

Due to the small sample size, there was no attempt to stratify participant demographic data such as age, gender, race, or whether they visited urban, rural, or inner-city radiology facilities.

Interview questions were framed in the context of the things that surprised, confused, or frustrated the patients in any of their experiences with radiology. This framework was selected as a result of previous focus group sessions held with patients on care experiences. Although partially overlapping, the three constructs were found to be sufficient to capture experiences related to care and administrative processes encountered by patients. Participants were encouraged to relate the questions to any experience, from the very first moment they were referred for radiology, to getting the results explained to them.

Specific questions asked of participants are shown in Table 1.

**Table 1.**
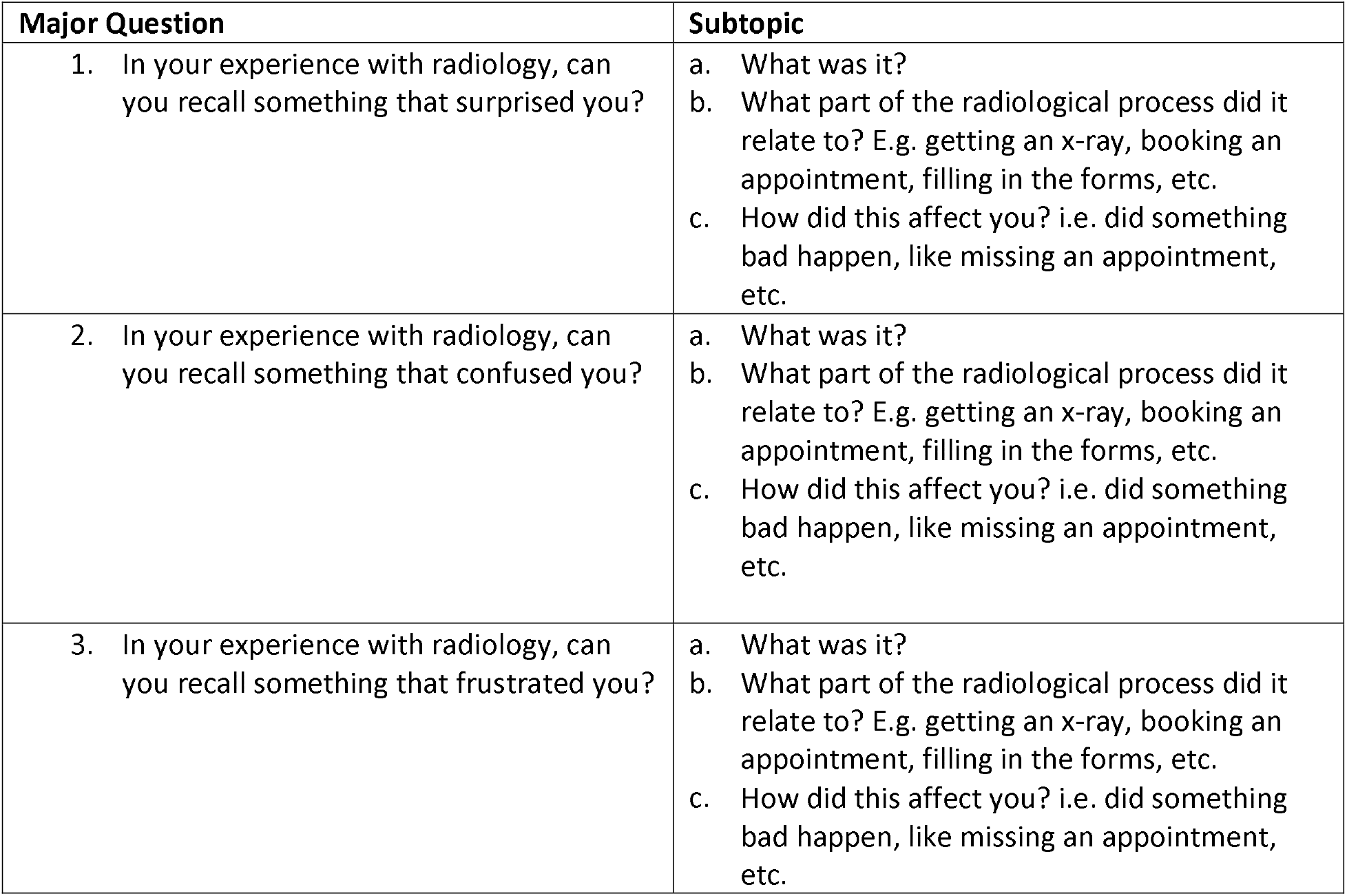
Question Matrix

The word cloud (Figure 1) was generated in the QDA from the text across all participants, and font size indicates word frequency, while color merely provides visual contrast.

Although the initial code structure matches the question elements, these elements were a vehicle needed for sufficiency of data collection, rather than a conceptual taxonomy for analysis, and were not used as part of the analytical process of constructing a code system. The code system was developed using a grounded theory approach. [6, 7] Using a card-sorting method, codes were grouped, split, or merged according to their affinity to each other, and through multiple iterative sorting sequences, and re-examination of the transcriptions or the associated audio tracks, four major code families were developed. The full code system is provided as a typology of radiology patient experiences in appendix A.

## Results

Communication was the most strongly discussed topic in the interview responses. The major area of ineffective communication related to processes; what the patient should expect from them, what the experience would be like, and what to expect after treatment.

While participants did report some concerns with care delivery, by far the majority of negative outcomes they reported related to the predictability and reliability of care and administrative processes. A complicating factor was that participants viewed the care journey in far broader terms than clinicians might expect, and experienced the bulk of their risks and issues with regard to weaknesses in care processes in the interstices. Where clinicians may think of the care journey starting at the unit door or even at first encounter with a clinician, participants were more likely to see the journey as starting with the motivating health event that led them to seek care, and ending only with resolution of a health condition.

There are indeed islands of excellence of well-executed process fragments that moved efficiently and expertly. Emergency Departments, for example, were related as being effective in maintaining process integrity. Once a patient had navigated themselves to the radiology department, the processes were described as generally working effectively. However, processes did not connect at the beginning or end with those outside the radiology department as robustly as desired.

The overall picture developed from analyzing the interview responses was of narrow and partially overlapping clinical domains with poor integration and frequent handoff and process failures, for which exhausted patients, were acting as the overall process managers.

Analysis of the interview text resulted in four major code families, namely “Patient Needs,” “Patient Psychological Security,” “Process,” and “Negative Outcomes.”

The “Patient Needs” code encompassed the accomodations that participants expressed regarding the mental and physical space in which care took place. Participants described physical barriers to accessing care, such as expensive or inconvenient parking, or facility layout, as well as needs such as care environment that is not noisy, cluttered, or smelly. A notable code was “Atypical Need”, which describes patient conditions that do not fit the standard care protocols well, resulting in care that was wasted, or that did not address the patient complaint.

**Table 2.** Patient Needs Code Family

| Code System | Code Frequency |
| --- | --- |
| **Needs** | (46) |
| Atypical Need | 6 |
| Care Environment | 12 |
| Disability | 7 |
| General Patient Needs | 11 |
| Parking | 4 |
| Physical Build | 6 |

Participants descibed needs and experiences that reflected what we termed “Psychological Security”. Several participants had mental health comorbidities that complicated the patient journey, and acted as barriers to care access or effective care use. Care environments that were low on “Compassion”, for example, acted as an access barrier, because participants were less motivated to engage with staff that seemed less compassionate. Likewise, a sense of low compassion reduced the efficacy of care provided, because participants attached less significance to advice from people they regarded as not compassionate. In comparison, participants who developed coping mechanisms or personal health networks had better success in overcoming obstacles.

**Table 3.** Patient Psychological Security

| Code System | Code Frequency |
| --- | --- |
| **Patient Psychological Security** | (26) |
| Comfort | 2 |
| Compassion | 8 |
| Coping Methods | 4 |
| Patient Confidence | 4 |
| Patient Health Literacy | 2 |
| Patient's Trust | 2 |
| Personal health network | 4 |

The “Process” code was the most active code family, and was threaded through almost every topic of discussion. Issues and missed opportunities related to poor communications featured strongly in how participants described their journey through radiology.

**Table 4.**
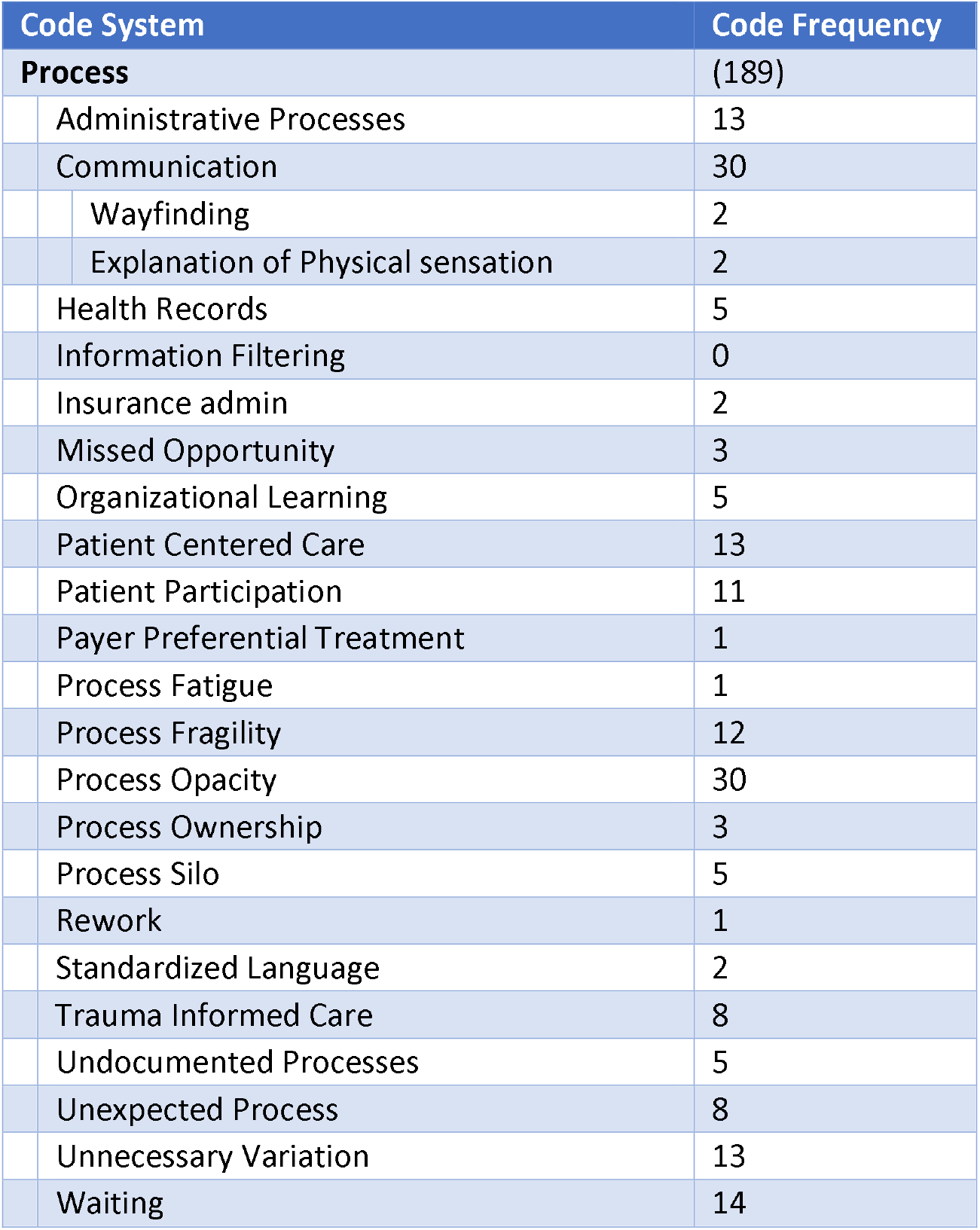
Care and Administration Processes

Finally, the major codes included the negative outcomes that participants encountered in their journeys through radiology. These codes reflected any negative experience or outcome, including any risks, issues, events, sequelae, or missed opportunity that the participants identified during their radiology journeys. Perhaps perculiar to radiology more than other specialties is the high frequency of “Scanxiety”, which denotes the anticipatory stress and worry resulting from the delay between having a radiology scan, and having the results explained and contextualized by a physician.

**Table 5.**
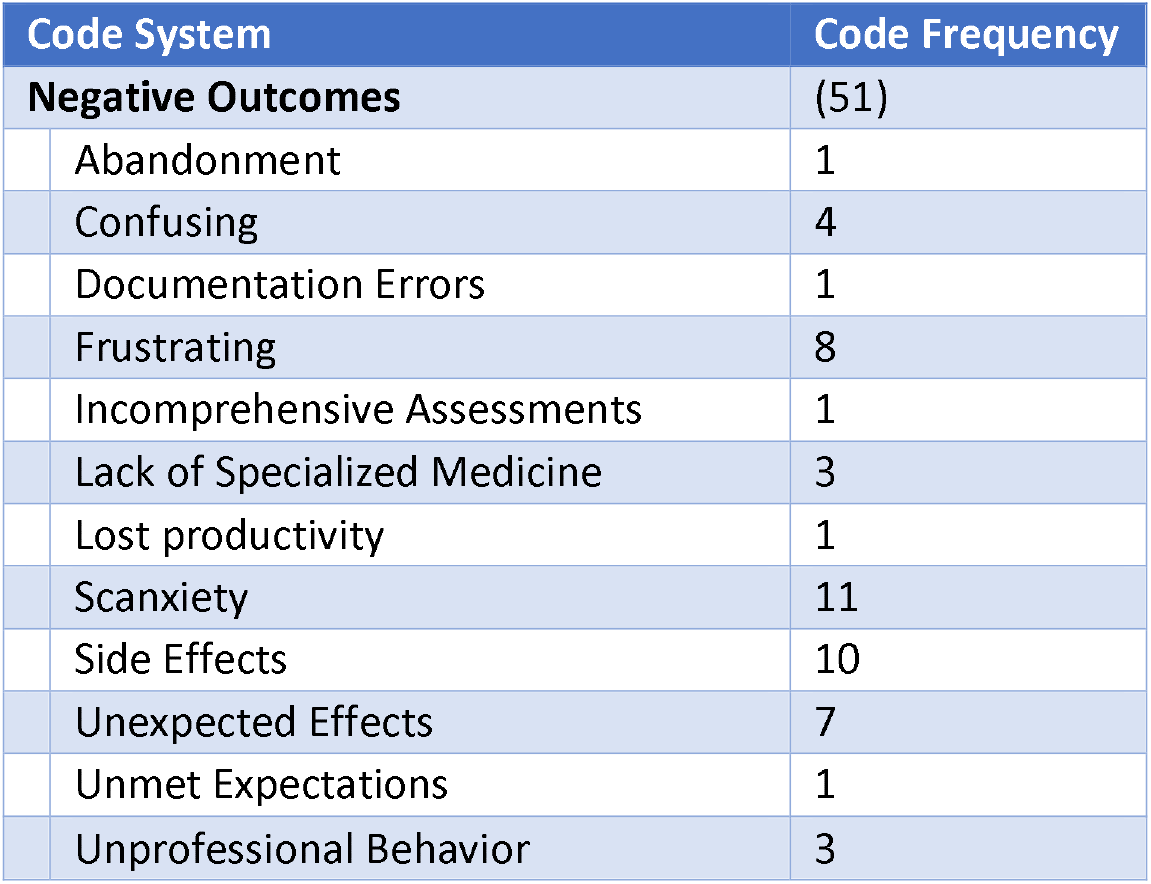
Negative Outcomes of the Care Experience

## Discussion

The radiology patient population has unique, yet reasonable needs and expectations. The care environment significantly influences the patient’s perception of quality and thus is a key determinant of how the patients may experience care.

### Process Failures

Participants spoke of fragile care processes that did not start as expected, frequently ground to a halt, or often failed to mesh well with the processes of other disciplines, unless the patient or their caregiver were actively monitoring and mitigating them. Participants were continuously acting as their own case manager, quality assessor, and process guide. Participants described how siloed processes resulted in negative outcomes such as being left alone over lunch breaks in an empty consultation room, aborted sessions due to ineffective coordination of care, or radiology teams performing scans that fit narrow protocols, but did not address the patient issues. In general, when processes failed, the patient was left carrying the burden of integrating or restarting them.

Participants reported rare occurrences where care processes within a specific radiology treatment or exam were at fault, but related persistent and ubiquitous process failures whenever processes between different specialties had to mesh, but failed to do so. Participants had exhaustive examples of inter-specialty process failures, whether those were radiology results never getting to the oncologist, physical therapist, or family doctor, or whether they were scheduling requests, referrals, or queries from myriad specialties never arriving intact in radiology. In some cases, participants waited for months for a radiology appointment that was never completely scheduled, or spent days trying to use the provider scheduling system, only to have to call and manually book an appointment.

Participants reported ongoing unease and uncertainty about whether appointments had been booked for the day, time, and location specified. Even when they were finally sure that a firm appointment was scheduled, these might be moved or cancelled by unknown parties without consultation. Likewise, even when patients went to extraordinary lengths to specify their special needs, they would often arrive only to find out that the service could not be completed as booked because the accommodations were not in place. For one participant, this meant costly and worrisome rescheduling for a dependent with special needs, and whose academic year dictated when lengthy radiology sessions could be scheduled.

Participants described being left alone in consultation rooms over lunch or having lights turned off in the examination room, or doors locked to the unit, because processes failed to mesh. In these cases, it was not that anyone was malicious or even negligent, but that the processes were simply fragile, poorly engineered, and gave no warning to anyone that they had failed.

Participants described how there was ultimately nobody other than themselves to take ownership of end-to-end processes, to report on broken processes, or to even notice that processes had failed.

### Unaddressed Patient Needs

Participants shared psychological needs including mental comfort and the confidence that processes and events will be predictable, be carried out with compassion, and uphold the trust that patients have in the clinicians, technicians, and administration staff they encounter.

Participants described physical needs that included sensory environment, seating location and ergonomics, and storage. Participants described the negative effects of having television news at high volume, unpleasantly high or low office temperatures, and inappropriate use of incense and other odorants. Participants described issues relating to lack of effective storage for hand luggage and personal items, and frequent lack of space for wheelchairs. Participants remarked on the benefits they experienced where reclining chairs and adequate lockable storage for personal baggage and effects was available.

Psychological needs were frequently unmet, and participants reported situations in which they felt confused or anxious. These needs included predictable wayfinding to reach the radiology unit, and predictable sensations, events, or interactions. Unexpected requirements for injections, confusing dietary restrictions, or difficulties in finding the radiology department undermined participants’ trust in their radiology teams. There was a strong sense that if simple expectations failed, the patient would have doubts about anything else the radiology team did - “if you cannot get this simple thing right, how can I trust anything else you tell me.”

Participants reported being bewildered by confusing or contradictory signage and lack of clear wayfinding markers to locate radiology departments. They were often surprised by sudden changes in their procedure preparations or unexpected sounds from equipment.

### Communication

Communication was the most discussed factor influencing the participant experience. Participants spoke of confusion, frustration, and surprise as result of insufficient preparatory discussion about what to expect, how it would feel, and what the possible post-treatment experience would be. In one example, the participant had expected the radiological treatment to “feel like something,” and the lack of any sensation of burning, tingling, or heat led them to doubt that the machine was on or working. In another example, confusing signage and presence of a television news crew for an event led the participant to be in a highly agitated state by the time they found the radiology department.

Nearly all of the participants referred to experiencing anxiety (“scanxiety”) during the waiting periods for receiving results. Participants felt that communications of their results, whether favorable or unfavorable, should be done as soon as feasible.

In some cases, participants expressed a perceived lack of full disclosure by providers about clinical findings. This resulted in feelings of distrust for some, and for others, reduced confidence in the competence of their provider. Participants felt disoriented by the failure of their providers to disclose the full range of possible side effects, especially likely long-term effects of treatment. Unexpected side effects were often associated with feelings of extreme anxiety and resulted in distrust of their providers. Although participants acknowledged that it may be cumbersome to run down the full list of possible side effects, they believed that the onus is on their care team to provide this information prior to discharge. Participants described side-effects and long-term effects not discussed with them, that ranged from inconvenient to debilitating.

Given the nature of radiological services, patients are often accustomed to certain procedural flows. Departures from the usual care procedures, without forewarning or explanation, was a concern for many of the participants. Many reported that changes were often unnecessary and only compounded the feeling of nervousness in an already stressful process. To mitigate their concerns, the participants felt that providers should 1) avoid departures from their usual approach if not clinically necessary and 2) discuss necessary changes in detail with the patient.

### Emotional Support

Participants spoke of a need for emotional support to be overtly demonstrated by provider staff, and participants reported often judging whether an encounter went well by how much compassion they experienced. Participants spoke of how comforting it was to interact with care teams that knew them by name, appeared to be concerned about their wellbeing, and expressed compassion when they needed reassurance. Many participants made specific mention of such experiences with the technicians, assistants, and administrative staff who consistently provided a personal touch. One participant expressed gratitude for being given a moment to cry due to being overwhelmed during an encounter.

In contrast, participants also reported a feeling of being rushed through treatments or scans without consideration for their psychological well-being and security. To elaborate, participants reported that some providers failed to talk them through the process or to ask about their comfort or their comprehension of the procedures in progress.

In addition, participants spoke of missed opportunities during visits. Providers may be focused strictly on the radiology aspects of an encounter, and not notice or react to signs that should drive up the index of suspicion. One participant was receiving radiological care that exposed ample evidence of severe domestic violence, but no one on the care team engaged the participant to see if they needed additional resources or were in an unsafe situation. The participant explained that they would have greatly benefited from the offer of additional support and resources at that time, and might have escaped an abusive situation far earlier.

### Coping Mechanisms for Unaddressed Patient Needs

To address the communications and emotional support challenges, some participants developed extensive coping mechanisms. Patients developed personal health networks (PHN), used mindfulness methods, built up their own health literacy, or scheduled encounters at times in the workday that enabled them to work for longer.

Participants reported leveraging social media and friendship circles to build PHNs. They used their PHNs to solicit assistance from radiologists, physicians, nurses, and technicians on social media in an effort to explain procedures, processes, or findings. These PHNs were effective in helping participants to self-manage their care journey, plan for encounters, or make sense of experiences or findings. While it may be laudatory that patients are taking control of their care in this manner, it also reveals gaps and risks in care delivery.

In one example, the instructions to drink no fluids prior to a scan were confusing, and the participant solicited help through Twitter physician connections to explain the restrictions in the context of the participant’s chronic condition. In another example, the participant used their social media connection with a radiologist to help understand the findings of a scan. In a further example, a participant exchanged spinal images with a radiologist in their PHN by sending a compact disk, and the PHN radiologist spent over an hour on the phone to discuss the images and their implications.

### Physical accommodations

Participants emphasized the importance of quick, seamless access to the care facility. This included conveniently located parking, direct access to the department of interest, accommodating waiting areas, and comfortable examination rooms. The layout of many waiting rooms often causes wheelchair-bound patients to be unable to sit with those accompanying them. Participants appreciated entertainment and informational resources tailored to patient needs, welcoming front desk staff, and settings that are calm, but were not in favor of television news or political programming in the waiting areas.

## Data Availability

Cleansed interview transcripts are available on reasonable request

## 1. Acknowledgments

We dedicate this report to Michelle Boyer, one of the participants for this study who passed away in the time since the conclusion of our interviews. Her insights into the radiology patient journey were both highly informative and deeply touching. We would also like to thank her parents who consented to this dedication and supported her engagement in this work.

